# Frequency and correlates of non-receipt of age-appropriate vaccination among children aged 6-35 months with medically attended diarrhea: Findings from the Enterics for Global Health (EFGH) *Shigella* study, 2022-2024

**DOI:** 10.64898/2025.12.04.25341661

**Authors:** Caren Oreso, Billy Ogwel, Alex O. Awuor, Raphael O. Anyango, Karen Kotloff, M. Jahangir Hossain, Henry Badji, Khuzwayo C. Jere, Latif Ndeketa, Josh Colston, Katia Manzanares Villanueva, Firdausi Qadri, Md Taufiqul Islam, Farah Naz Qamar, Naveed Ahmed, Sonia I. Rao, Patricia B. Pavlinac, Richard Omore

## Abstract

**Background:** Complete childhood immunization protects children from long-term health complications and disabilities caused by vaccine-preventable diseases. Enterics for Global Health (EFGH)-*Shigella* surveillance was a two-year study measuring incidence rates and consequences of *Shigella* among children aged 6-35 months in seven sites located in Asia, Latin America and Africa. Here, we estimated the prevalence and factors associated with non-receipt of age-appropriate vaccination among children enrolled in the EFGH-*Shigella* study.

**Methods:** In this nested cross-sectional study, we analysed data from 7,932 children aged 6-35 months presenting with medically attended diarrhea (MAD). Vaccines recommended per each country’s national immunization schedule were extracted from medical records and risk factors were collected by caregiver interview and physical exam. We defined age-appropriate vaccines as receipt of the early childhood vaccinations within one month of the recommended age, according to the national immunization schedule, on the immunization card. Poison regression was used to identify independent factors associated with non-receipt of age-appropriate vaccination accounting for all covariates.

**Results:** Over half of enrolled children (51.7%) did not receive all age-appropriate vaccines most commonly in The Gambia (75.2%) and least frequently in Bangladesh (22.3%). Children 12-35 months of age were more likely not have all age appropriate vaccines compared to children 6-11 months (aPR: 1.47, 95%CI 1.39 to 1.54), children who came from households with ≥3 children aged <5 years (aPR:1.07;1.01-1.13), had mothers with low education (aPR:1.18; 1.12-1.25), and were wasted (Moderately: aPR: 1.06; 1.00-1.13; Severely: aPR : 1.14, 1.04-1.25) were more likely to miss all age-appropriate vaccines compared to their counterparts who did not.

**Conclusion:** Non-receipt of age-appropriate vaccination was largely age dependent, driven by mother’s education and severe wasting highlighting the need to design effective strategies that incorporate site complexities to improve timely vaccination targeting vulnerable groups.

## Introduction

Infectious diseases are the major cause of morbidity and mortality in low-and-middle-income countries (LMICs) [1]. Globally, in 2019, the proportion of disability-adjusted life-years (DALYs) associated with potential vaccine preventable infectious diseases in children under-5 years was highest in sub-Saharan Africa, South Asia and Latin America [2]; the same places with the lowest vaccine uptake [3]. In 2019, vaccine preventable deaths from diarrhea alone accounted for nearly ten percent of all under-5 deaths [1].

Immunization is a proven lifesaving intervention and several vaccines exist for infectious diseases such as rotavirus, measles, tuberculosis, tetanus, diphtheria, polio, and pertussis. However, despite the preventability of these diseases and universal availability of early childhood vaccinations, these conditions continue to be the main drivers of morbidity and mortality in LMICs [2]. In 2023, two years after the World Health Organization (WHO) kicked off the ambitious new Immunization Agenda 2030 (IA2030), 40.8% of children did not receive any diphtheria, pertussis, and tetanus vaccinations [4]. Few studies have quantitatively examined age-appropriate vaccination in LMICs and those that have, are often limited by small sample sizes and focused on single-country contexts [5–7]. By leveraging data from the Enterics for Global Health (EFGH) *Shigella* surveillance study—which spans countries across Africa, South Asia, and Latin America—our study provides an added advantage of a large, diverse sample that provides broader insights across multiple settings with demand for vaccination against infectious diseases. Understanding factors of non-receipt of age appropriate vaccination in LMICs could help inform resource allocation and strategies aimed at optimising vaccine uptake, and help achieve child health targets.

EFGH study is a two-year study designed to establish baseline incidence rates and consequences of *Shigella* diarrhea with the intent to inform *Shigella* vaccine trials and introduction in countries with the highest disease burden [8]. Here, we estimated prevalence and factors associated with non-receipt of age-appropriate vaccination among children aged 6-35 months enrolled with medically attended diarrhea (MAD) in the EFGH study.

## Methods

### Study setting and Population

The EFGH study sites are located in Bangladesh, Kenya, Malawi, Mali, Pakistan, Peru and The Gambia and the study was conducted in populations that represent high infectious and diarrheal disease burden settings on the three continents. Detailed methods [8] along with characteristics of the study settings and populations are described in detail elsewhere [9–15].

### Definitions

*Vaccine completeness* included assessing vaccine receipt and timing of the following vaccines: Bacillus Calmette-Guérin (BCG), Polio, DPT (Diphtheria, Pertussis and Tetanus) Hepatitis B, *Haemophilus influenzae* type b (Hib), Rotavirus, MMR (Measles+Mumps+Rubella) and Pneumococcal conjugate vaccine (PCV). Rotavirus vaccination has not been introduced into the national immunization schedule in Bangladesh therefore considered vaccines receipt as above except Rotavirus to meet the complete vaccine definition in Bangladesh.

*Non-receipt of age-appropriate vaccination* was defined as absence of a record of having received the eight (or seven for Bangladesh) vaccinations within one month of the recommended age, according to the national immunization schedule, on the immunization card.

*Diarrhea* was defined as a caregiver report of three or more abnormally loose or watery stools in the previous 24 hours.

*Medically attended diarrhea (MAD)* was defined as diarrhea that led to seeking care at a designated EFGH study sentinel facility by the caregiver.

### Study design

This was a secondary, cross-sectional analysis of baseline collected in the EFGH study.

#### Inclusion criteria

Children with MAD aged 6-35 months, residing in a demographically defined study catchment population, who sought care at a designated study sentinel health facility with a vaccination card, planned to remain at their current residence for at least 4 months, and whose primary caregiver was able to provide written informed consent, were included in this analysis.

#### Exclusion criteria

Children were excluded if younger than 6 months or older than 35 months and presented with non-diarrhea conditions, if the caregiver-reported diarrhea did not meet the study definition, if the primary caregiver did not provide written informed consent or did not reside in the study catchment area, if vaccination cards were not available, if four or more hours passed between when the child presented at the health facility and the study screening occurred, if the site enrolment sample size had been met, or if child was referred to a non-study facility for further treatment.

### Data Collection

The information collected included basic patient and family demographics, geographic, socio-economic and clinical characteristics, water and sanitation variables as described elsewhere [16]. Vaccination information was abstracted from the immunization cards.

### Statistical Analysis

Frequencies and percentages were used to summarize the vaccination status of children with reported MAD. Overall and site-specific estimates for each vaccine and overall age-appropriate vaccination were calculated. We employed descriptive statistics to summarize factors associated with non-receipt of age-appropriate vaccination. Continuous variables were presented using median and interquartile range, and comparisons between groups were performed using the Wilcoxon Rank-Sum test. Categorical variables were summarized using frequencies and proportions, and group comparisons were conducted using the Chi-square test.

Poisson regression was used to identify factors associated with non-receipt of age-appropriate vaccination; the main binary outcome of interest which has been described previously [17]. Initially, all potential predictor factors were assessed in separate individual bivariate Poisson regression models. Variables with a p-value < 0.2 were selected for inclusion in the multivariable model. The crude and adjusted prevalence ratios (aPR) were computed with 95% confidence intervals (95% CI). We also conducted sensitivity analyses, stratifying the models by age and site, separately.

### Ethical Considerations

This analysis involved data collected as part of EFGH protocol which was approved by the respective IRBs at all the country sites prior to study initiation [8]. All study participants provided written informed consent [8].

## Results

From June 2022 to August 2024, we screened 30,191 children with MAD; of whom 9,476 (31.4%) were enrolled in the EFGH study (Fig 1).

**Figure 1.**
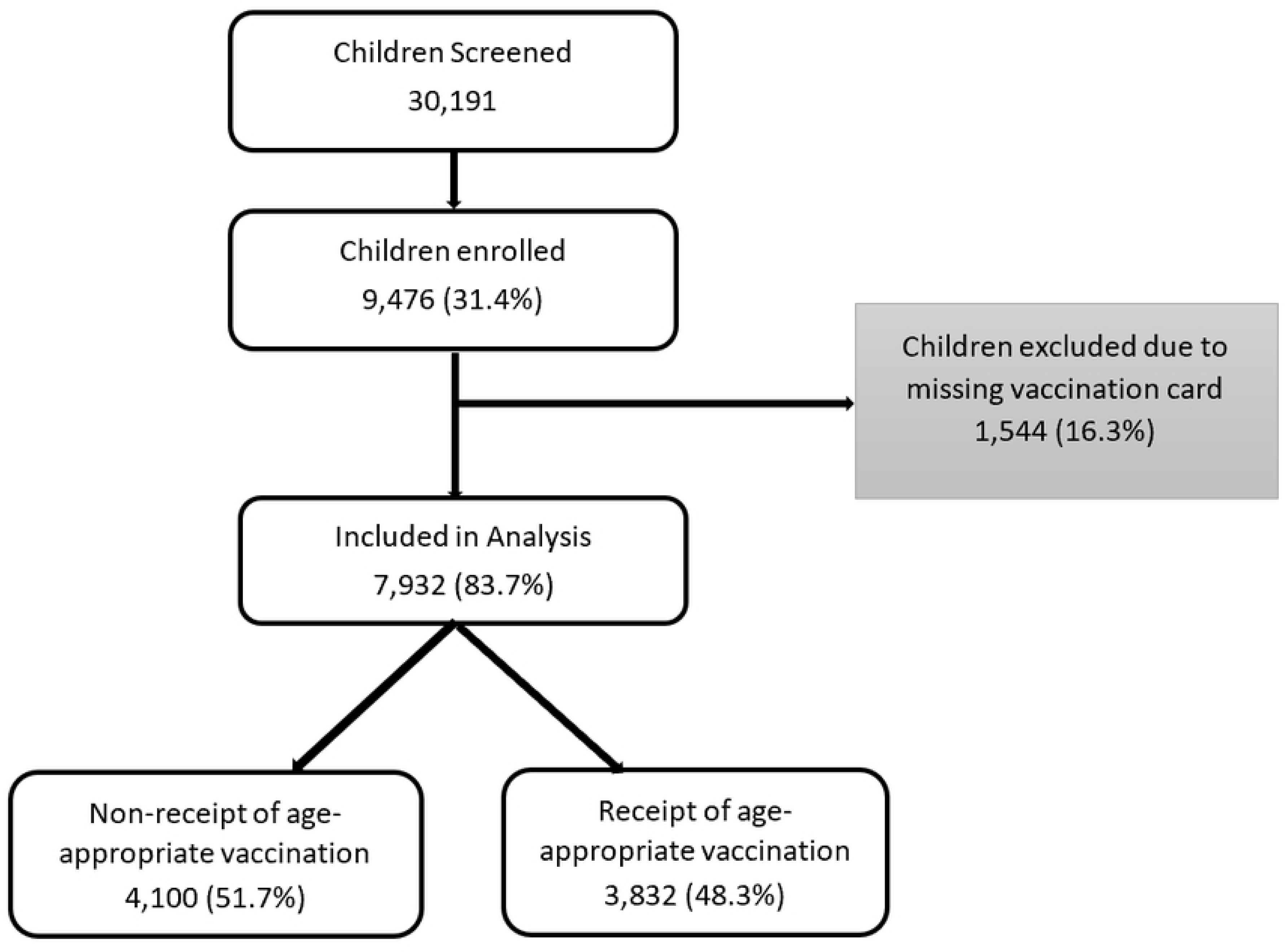
Flowchart of enrolment of children aged 6-35 months presenting with medically attended diarrhea and vaccination status, EFGH *Shigella* surveillance study, June 2022 - August 2024

We excluded 1,544 (16.3%) children in this secondary analysis due to missing a vaccine card. Of the 7,932 children included, the median (interquartile range [IQR]) age of the children analysed was 14 (9-21) months and 4,296 (54.2%) children were male. The caregiver’s median (IQR) age was 25 (22-30) years and 52.2% of these were mothers who had low education (primary school only or no education). Approximately 22% of children were stunted. Majority (76.0 %) of caregivers were not working at the time the child was enrolled in this study. Among the children enrolled 4,617 (58.2%), 1,385 (17.5%), 1,930 (24.3%) had mild, moderate and severe diarrhea, respectively (Table 1).

**Table 1:** Characteristics of study population stratified by vaccination status, EFGH *Shigella* surveillance study, June 2022 - August 2024.

| Variable | Category | Overall<br>(N=7,932) | Non-receipt of Age<br>appropriate<br>Vaccination<br>(N=4,100, 51.7%) | Age-appropriate<br>Vaccination<br>(N=3,832, 48.3%) |
| --- | --- | --- | --- | --- |
| **Demographic characteristics** |  | **n (%)** | **n (%)** | **n (%)** |
| Age in months | Median [Q1-Q3] | 14 [9-21] | 16 [10-22] | 12 [8-16] |
| Age category | 6-11m | 3167 (39.9) | 1264 (30.8) | 1903 (49.7) |
|  | 12-17m | 2145 (27.0) | 1026 (25.0) | 1119 (29.2) |
|  | 18-23m | 1454 (18.3) | 935 (22.8) | 519 (13.5) |
|  | 24-35m | 1166 (14.7) | 875 (21.3) | 291 (7.6) |
| Sex | Male | 4296 (54.2) | 2189 (53.4) | 2107 (55.0) |
| Site | Bangladesh | 1211 (15.3) | 270 (6.6) | 941 (24.6) |
|  | Kenya | 1211 (15.3) | 867 (21.1) | 344 (9.0) |
|  | Malawi | 954 (12.0) | 542 (13.2) | 412 (10.8) |
|  | Mali | 1229 (15.5) | 506 (12.3) | 723 (18.9) |
|  | Pakistan | 992 (12.5) | 527 (12.9) | 465 (12.1) |
|  | Peru | 946 (11.9) | 344 (8.4) | 602 (15.7) |
|  | The Gambia | 1389 (17.5) | 1044 (25.5) | 345 (9.0) |
| No. of children <5 yrs in household | <3<br>≥3 | 5663 (71.4)<br>2269 (28.6) | 2718 (66.3)<br>1382 (33.7) | 2945 (76.9)<br>887 (23.1) |
| **Socio-economic characteristics** |  |  |  |  |
| Caregiver age in years | Median [Q1-Q3] | 25 [22-30] | 26 [22-30] | 25 [22-30] |
| Final Quintile | Quintile 1 - Least wealthy | 1238 (15.6) | 717 (17.5) | 521 (13.6) |
|  | Quintile 2 | 2252 (28.4) | 1256 (30.6) | 996 (26) |
|  | Quintile 3 | 2158 (27.2) | 1120 (27.3) | 1038 (27.1) |
|  | Quintile 4 | 1498 (18.9) | 755 (18.4) | 743 (19.4) |
|  | Quintile 5 - Wealthiest | 786 (9.9) | 252 (6.1) | 534 (13.9) |
| Mother's education<sup>β</sup> | High Education | 3782 (47.8) | 1650 (40.3) | 2132 (55.7) |
|  | Low Education | 4137 (52.2) | 2442 (59.7) | 1695 (44.3) |
| Father's education<sup>β</sup> | High Education | 4190 (56) | 1951 (51) | 2239 (61.2) |
|  | Low Education | 3293 (44) | 1874 (49) | 1419 (38.8) |
| Caregiver occupation<sup>¥</sup> | No employment | 5977 (76) | 2855 (70.1) | 3122 (82.2) |
|  | Formal employment | 143 (1.8) | 57 (1.4) | 86 (2.3) |
|  | Informal employment | 1747 (22.2) | 1158 (28.5) | 589 (15.5) |
| **Clinical characteristics** |  |  |  |  |
| Diarrhea duration in days | Median [Q1-Q3] | 4 [3-6] | 4 [3-6] | 4 [3-6] |
| Respiratory rate * | Normal | 6980 (88) | 3632 (88.6) | 3348 (87.4) |
|  | Low | 562 (7.1) | 208 (5.1) | 354 (9.2) |
|  | High | 390 (4.9) | 260 (6.3) | 130 (3.4) |
| Heart rate * | Normal | 7198 (91.7) | 3775 (93.8) | 3423 (89.5) |
|  | Low | 644 (8.2) | 243 (6) | 401 (10.5) |
|  | High | 7 (0.1) | 6 (0.1) | 1 (0) |
| Dysentery | Yes | 1033 (13) | 576 (14) | 457 (11.9) |
| Prolonged<sup>£</sup> | Yes | 1518 (19.1) | 646 (15.8) | 872 (22.8) |
| Persistent<sup>£</sup> | Yes | 174 (2.2) | 69 (1.7) | 105 (2.7) |
| Dehydration | No sign of Dehydration | 5957 (75.1) | 3061 (74.7) | 2896 (75.6) |
|  | Severe | 97 (1.2) | 74 (1.8) | 23 (0.6) |
|  | Some | 1878 (23.7) | 965 (23.5) | 913 (23.8) |
| Any subsequent episodes | Yes | 2680 (33.8) | 1334 (32.5) | 1346 (35.1) |
| Modified Vesikari Score | Mild | 4617 (58.2) | 2436 (59.4) | 2181 (56.9) |
|  | Moderate | 1385 (17.5) | 715 (17.4) | 670 (17.5) |
|  | Severe | 1930 (24.3) | 949 (23.1) | 981 (25.6) |
| Stunted | No | 6167 (78.2) | 3094 (75.8) | 3073 (80.7) |
|  | Yes | 1724 (21.8) | 989 (24.2) | 735 (19.3) |
| Wasted | None | 6545 (82.9) | 3318 (81.2) | 3227 (84.7) |
|  | Moderate | 1061 (13.4) | 585 (14.3) | 476 (12.5) |
|  | Severe | 290 (3.7) | 181 (4.4) | 109 (2.9) |
| Prior care-seeking | Yes | 1707 (21.5) | 646 (15.8) | 1061 (27.7) |
| Seeking care in the future | Easy | 7156 (90.8) | 3641 (89.3) | 3515 (92.5) |
|  | Challenging | 724 (9.2) | 437 (10.7) | 287 (7.5) |
| Accept new vaccine | Never | 83 (1.1) | 43 (1.1) | 40 (1.1) |
|  | Sometimes | 440 (5.7) | 275 (6.9) | 165 (4.4) |
|  | Yes | 7216 (93.2) | 3680 (92) | 3536 (94.5) |
| Drinking water | Improved | 7423 (93.6) | 3761 (91.7) | 3662 (95.6) |
|  | Unimproved | 509 (6.4) | 339 (8.3) | 170 (4.4) |
| Sanitation | Improved | 6517 (82.2) | 3215 (78.4) | 3302 (86.2) |
|  | Unimproved | 1415 (17.8) | 885 (21.6) | 530 (13.8) |
| Breastfeeding | Yes | 476 (6) | 264 (6.5) | 212 (5.6) |

<sup>β</sup> High education- some secondary, and secondary school or greater; Low education: No education, less than primary, primary school only, and Koranic school only.
<sup>¥</sup> No employment-Not employed, housewife, and student; Formal employment- professional; Informal employment-business (self-employed), business (other employer), casual laborer, and farmer.
\*Cutoffs for heart rate and respiratory rate based on the Pediatric Advanced Life Support (PALS) guidelines.
<sup>£</sup> ≥7 days of diarrhea during index diarrhea episode
<sup>£</sup> ≥14 days of diarrhea during index diarrhea episode

### Prevalence of non-receipt of age-appropriate vaccination

Among the 7,932 children included, 4,100 (51.7%) had not received all age-appropriate vaccinations at the time of enrolment (Fig. 1). In general, the prevalence of non-receipt of age-appropriate vaccination varied across the study sites with The Gambia reporting the highest (75.2%) and Bangladesh the lowest prevalence (22.3%) (Table 2). Overall, the three vaccines with the highest prevalence of age-appropriate non-receipt were polio, MMR and PCV at 26.8%,19.3% and 14.1%, respectively. The vaccine with the lowest non-receipt of age appropriate vaccination uptake across all sites was BCG (2.5%).

**Table 2:**
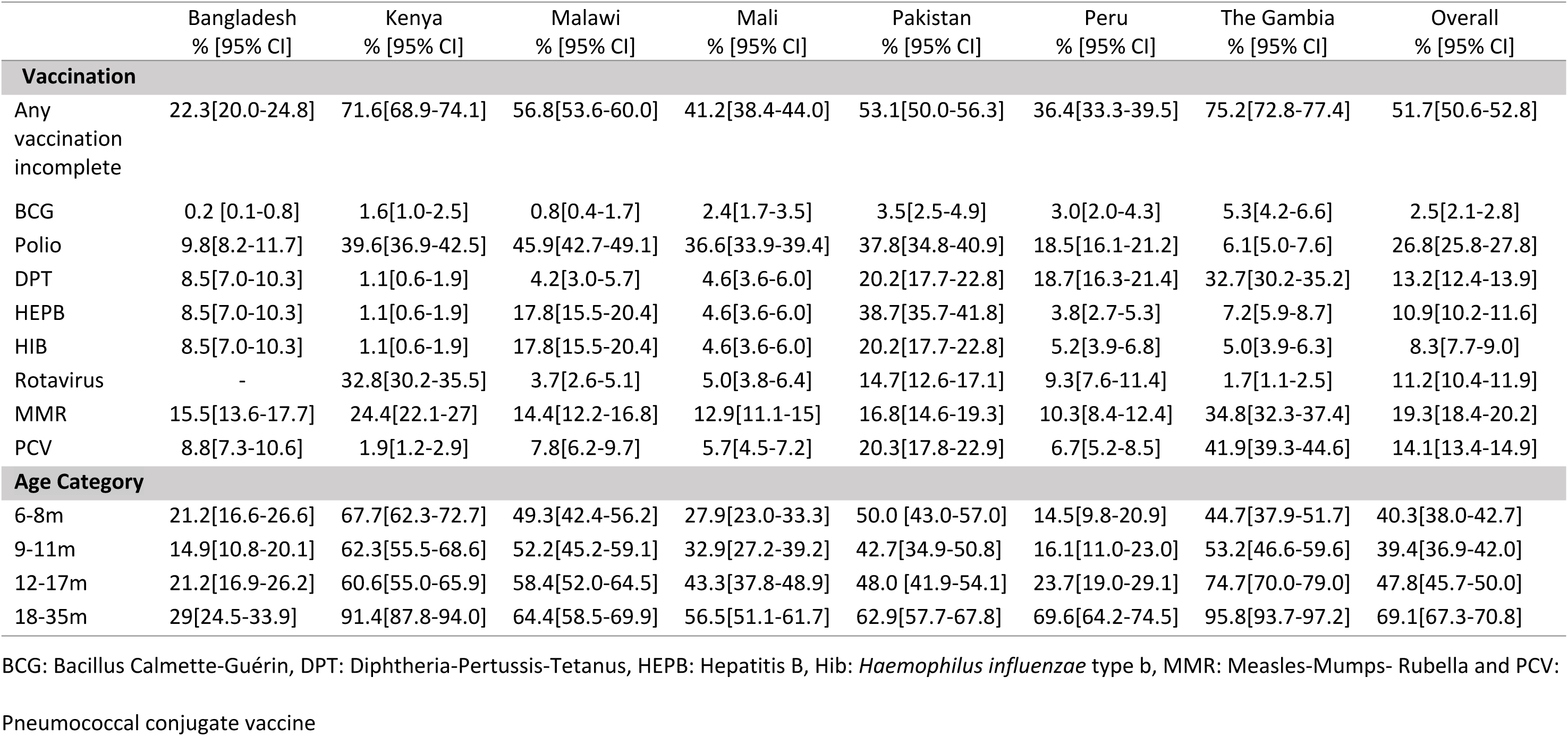
Proportion of children 6-35 months presenting with medically-attended diarrhea who had non-receipt of age-appropriate vaccination, by vaccine and age stratified by sites, EFGH *Shigella* surveillance study, June 2022 - August 2024.

|  | Bangladesh<br>% [95% CI] | Kenya<br>% [95% CI] | Malawi<br>% [95% CI] | Mali<br>% [95% CI] | Pakistan<br>% [95% CI] | Peru<br>% [95% CI] | The Gambia<br>% [95% CI] | Overall<br>% [95% CI] |
| --- | --- | --- | --- | --- | --- | --- | --- | --- |
| **Vaccination** |  |  |  |  |  |  |  |  |
| Any vaccination incomplete | 22.3[20.0-24.8] | 71.6[68.9-74.1] | 56.8[53.6-60.0] | 41.2[38.4-44.0] | 53.1[50.0-56.3] | 36.4[33.3-39.5] | 75.2[72.8-77.4] | 51.7[50.6-52.8] |
| BCG | 0.2 [0.1-0.8] | 1.6[1.0-2.5] | 0.8[0.4-1.7] | 2.4[1.7-3.5] | 3.5[2.5-4.9] | 3.0[2.0-4.3] | 5.3[4.2-6.6] | 2.5[2.1-2.8] |
| Polio | 9.8[8.2-11.7] | 39.6[36.9-42.5] | 45.9[42.7-49.1] | 36.6[33.9-39.4] | 37.8[34.8-40.9] | 18.5[16.1-21.2] | 6.1[5.0-7.6] | 26.8[25.8-27.8] |
| DPT | 8.5[7.0-10.3] | 1.1[0.6-1.9] | 4.2[3.0-5.7] | 4.6[3.6-6.0] | 20.2[17.7-22.8] | 18.7[16.3-21.4] | 32.7[30.2-35.2] | 13.2[12.4-13.9] |
| HEPB | 8.5[7.0-10.3] | 1.1[0.6-1.9] | 17.8[15.5-20.4] | 4.6[3.6-6.0] | 38.7[35.7-41.8] | 3.8[2.7-5.3] | 7.2[5.9-8.7] | 10.9[10.2-11.6] |
| HIB | 8.5[7.0-10.3] | 1.1[0.6-1.9] | 17.8[15.5-20.4] | 4.6[3.6-6.0] | 20.2[17.7-22.8] | 5.2[3.9-6.8] | 5.0[3.9-6.3] | 8.3[7.7-9.0] |
| Rotavirus | - | 32.8[30.2-35.5] | 3.7[2.6-5.1] | 5.0[3.8-6.4] | 14.7[12.6-17.1] | 9.3[7.6-11.4] | 1.7[1.1-2.5] | 11.2[10.4-11.9] |
| MMR | 15.5[13.6-17.7] | 24.4[22.1-27] | 14.4[12.2-16.8] | 12.9[11.1-15] | 16.8[14.6-19.3] | 10.3[8.4-12.4] | 34.8[32.3-37.4] | 19.3[18.4-20.2] |
| PCV | 8.8[7.3-10.6] | 1.9[1.2-2.9] | 7.8[6.2-9.7] | 5.7[4.5-7.2] | 20.3[17.8-22.9] | 6.7[5.2-8.5] | 41.9[39.3-44.6] | 14.1[13.4-14.9] |
| **Age Category** |  |  |  |  |  |  |  |  |
| 6-8m | 21.2[16.6-26.6] | 67.7[62.3-72.7] | 49.3[42.4-56.2] | 27.9[23.0-33.3] | 50.0 [43.0-57.0] | 14.5[9.8-20.9] | 44.7[37.9-51.7] | 40.3[38.0-42.7] |
| 9-11m | 14.9[10.8-20.1] | 62.3[55.5-68.6] | 52.2[45.2-59.1] | 32.9[27.2-39.2] | 42.7[34.9-50.8] | 16.1[11.0-23.0] | 53.2[46.6-59.6] | 39.4[36.9-42.0] |
| 12-17m | 21.2[16.9-26.2] | 60.6[55.0-65.9] | 58.4[52.0-64.5] | 43.3[37.8-48.9] | 48.0 [41.9-54.1] | 23.7[19.0-29.1] | 74.7[70.0-79.0] | 47.8[45.7-50.0] |
| 18-35m | 29[24.5-33.9] | 91.4[87.8-94.0] | 64.4[58.5-69.9] | 56.5[51.1-61.7] | 62.9[57.7-67.8] | 69.6[64.2-74.5] | 95.8[93.7-97.2] | 69.1[67.3-70.8] |

BCG: Bacillus Calmette-Guérin, DPT: Diphtheria-Pertussis-Tetanus, HEPB: Hepatitis B, Hib: *Haemophilus influenzae* type b, MMR: Measles-Mumps- Rubella and PCV:
Pneumococcal conjugate vaccine

### Factors associated with non-receipt of age-appropriate vaccination

Compared to children aged 6 to 11 months of age, children 12 months and above had a higher likelihood of missing all age-appropriate vaccines (Table 3). Similarly, compared to Kenya, all other EFGH country sites were less likely to have children missing age-appropriate vaccines with The Gambia (adjusted Prevalence Ratio [aPR]:0.91, [95%CI: 0.83-0.99]) and Malawi (aPR: 0.78; 0.72-0.85) being the least likely to have children missing vaccines. Additionally, children who came from households with ≥3 children aged <5 years (aPR:1.07;1.01-1.13), had mothers with low education (aPR:1.18; 1.12-1.25), and were wasted (Moderately: aPR: 1.06; 1.00-1.13; Severely: aPR : 1.14, 1.04-1.25) were more likely to miss all age-appropriate vaccines compared to their counterparts who did not (Table 3).

**Table 3:** Correlates of non-receipt of age-appropriate vaccination among children aged 6-35 months presenting with medically-attended diarrhea, EFGH *Shigella* surveillance study, June2022 - August 2024.

| Variable | Category | Bivariate |  | Multivariable |  |
| --- | --- | --- | --- | --- | --- |
|  |  | Unadjusted Prevalence Ratio [95% CI] | P-value | Adjusted Prevalence Ratio [95% CI] | P-value |
| Age category | 6-11m | Ref |  | Ref |  |
|  | 12-17m | **1.20 [1.13-1.27]** | **<0.001** | **1.21 [1.13-1.29]** | **<0.001** |
|  | 18-23m | **1.61 [1.52-1.71]** | **<0.001** | **1.56 [1.47-1.65]** | **<0.001** |
|  | 24-35m | **1.88 [1.78-1.98]** | **<0.001** | **1.85 [1.75-1.96]** | **<0.001** |
| Sex | Male | 0.97 [0.93-1.01] | 0.154 | 0.97 [0.93-1.01] | 0.163 |
| Site | Kenya | Ref |  | Ref |  |
|  | Bangladesh | **0.31 [0.28-0.35]** | **<0.001** | **0.32 [0.28-0.36]** | **<0.001** |
|  | Malawi | **0.79 [0.74-0.85]** | **<0.001** | **0.78 [0.72-0.85]** | **<0.001** |
|  | Mali | **0.58 [0.53-0.62]** | **<0.001** | **0.53 [0.47-0.58]** | **<0.001** |
|  | Pakistan | **0.74 [0.69-0.79]** | **<0.001** | **0.65 [0.59-0.71]** | **<0.001** |
|  | Peru | **0.51 [0.46-0.56]** | **<0.001** | **0.51 [0.46-0.57]** | **<0.001** |
|  | The Gambia | **1.05 [1.00-1.10]** | **0.041** | **0.91 [0.83-0.99]** | **0.021** |
| Number of children < 5 years | 1-2 | Ref |  | Ref |  |
| | $\geq 3$ | **1.27 [1.22-1.32]** | **<0.001** | **1.07 [1.01-1.13]** | **0.023** |
| Caregiver age in years |  | 1.00 [0.99-1.01] | 0.068 | **1.00 [1.00-1.00]** | **0.019** |
| Wealth Quintile | Quintile 1-Least wealthy | Ref |  | Ref |  |
|  | Quintile 2 | 0.96 [0.91-1.02] | 0.218 | **0.93 [0.87-0.99]** | **0.016** |
|  | Quintile 3 | **0.9 [0.84-0.95]** | **0.001** | **0.92 [0.86-0.98]** | **0.010** |
|  | Quintile 4 | 0.87 [0.81-0.93] | **<0.001** | 0.98 [0.90-1.05] | 0.535 |
|  | Quintile 5-Wealthiest | **0.55 [0.49-0.62]** | **<0.001** | **0.88 [0.77-0.99]** | **0.037** |
| Mother<br>education<sup>β</sup> | Low Education | **1.35 [1.29-1.41]** | **<0.001** | **1.18 [1.12-1.25]** | **<0.001** |
| Father<br>education<sup>β</sup> | Low Education | **1.22 [1.17-1.28]** | **<0.001** | **1.06 [1.00-1.11]** | **0.032** |
| Caregiver<br>occupation* | No<br>employment | Ref |  | Ref |  |
|  | Formal<br>employment | 0.83 [0.68-1.02] | 0.081 | 0.91 [0.75-1.10] | 0.329 |
|  | Informal<br>employment | **1.39 [1.33-1.45]** | **<0.001** | 1.01 [0.97-1.06] | 0.609 |
| Diarrhea<br>duration in days |  | **0.97 [0.96-0.97]** | **<0.001** | 1.00 [0.99-1.01] | 0.847 |
| Respiratory<br>rate* | Normal | Ref |  | Ref |  |
|  | Low | **0.71 [0.64-0.79]** | **<0.001** | 0.92 [0.82-1.02] | 0.127 |
|  | High | **1.28 [1.19-1.38]** | **<0.001** | 1.06 [0.98-1.16] | 0.167 |
| Heart rate* | Normal | Ref |  | Ref |  |
|  | Low | **0.72 [0.65-0.8]** | **<0.001** | 0.90 [0.81-1.00] | 0.060 |
|  | High | **1.63 [1.21-2.21]** | **0.001** | 1.33 [0.89-2.00] | 0.163 |
| Dysentery | Yes | **1.09 [1.03-1.16]** | **0.004** | 1.04 [0.99-1.1] | 0.144 |
| Prolonged<sup>£</sup> | Yes | **0.79 [0.74-0.84]** | **<0.001** | 1.04 [0.95-1.13] | 0.421 |
| Persistent<sup>£</sup> | Yes | **0.76 [0.63-0.92]** | **0.004** | 1.01 [0.80-1.26] | 0.956 |
| Dehydration<sup>£</sup> | None | Ref |  | Ref |  |
|  | Severe | **1.48 [1.33-1.66]** | **<0.001** | - | - |
|  | Some | 1.00 [0.95-1.05] | 1.000 | - | - |
| Any subsequent<br>episodes | Yes | **0.95 [0.9-0.99]** | **0.016** | **1.05 [1.00-1.1]** | **0.049** |
| Modified<br>Vesikari Score | Mild | Ref |  | Ref |  |
|  | Moderate | 0.98 [0.92-1.04] | 0.460 | 0.99 [0.94-1.05] | 0.763 |
|  | Severe | **0.93 [0.88-0.98]** | **0.009** | 1.03 [0.98-1.09] | 0.226 |
| Stunted | Yes | **1.14 [1.09-1.20]** | **<0.001** | 1.00 [0.96-1.06] | 0.865 |
| Wasted | None | Ref |  | Ref |  |
|  | Moderate | **1.09 [1.02-1.15]** | **0.006** | **1.06 [1.00-1.13]** | **0.045** |
|  | Severe | **1.23 [1.12-1.35]** | **<0.001** | **1.14 [1.04-1.25]** | **0.006** |
| Prior care-seeking | Yes | 0.68 [0.64-0.73] | **<0.001** | 1.00 [0.93-1.07] | 1.000 |
| seeking care in<br>the future | Challenging | **1.19 [1.11-1.26]** | **<0.001** | **0.88 [0.82-0.94]** | **<0.001** |
| Accept new<br>vaccine | Never | Ref |  | Ref |  |
|  | Sometimes | 1.21 [0.97-1.5] | 0.094 | 1.10 [0.89-1.36] | 0.385 |
|  | Yes | 0.98 [0.8-1.21] | 0.882 | 0.90 [0.74-1.10] | 0.310 |
| Drinking water | Improved | Ref |  | Ref |  |
|  | Unimproved | **1.31 [1.23-1.40]** | **<0.001** | 1.00 [0.92-1.08] | 0.920 |
| Sanitation | Improved | Ref |  | Ref |  |
|  | Unimproved | **1.27 [1.21-1.33]** | **<0.001** | 1.00 [0.95-1.05] | 0.988 |
| breastfeeding | Yes | 1.08 [0.99-1.17] | 0.079 | 1.05 [0.97-1.14] | 0.244 |

<sup>B</sup>- High education- some secondary, and secondary school or greater; Low education: No education, less than primary, primary school only, and Koranic school only.
<sup>¥</sup>- No employment- Not employed, housewife, and student; Formal employment- professional; Informal employment-business (self-employed), business (other employer), casual laborer, and farmer.
\*Cutoffs for heart rate and respiratory rate based on the Pediatric Advanced Life Support (PALS) guidelines.
<sup>£</sup>≥7 days of diarrhea during index diarrhea episode
<sup>£</sup>≥14 days of diarrhea during index diarrhea episode
<sup>‡</sup> Dehydration excluded from multivariable model as it's a component of Vesikari score.

Conversely, children who came from wealthier households (quintile 5) compared to least wealthy households (aPR: 0.88; 0.77-0.99) were less likely to miss all age-appropriate vaccinations. Similarly, children whose caregivers reported finding it challenging to seek care in the future compared to those who did not [aPR: 0.88 [0.82-0.94]] were less likely to miss all age-appropriate vaccinations (Table 3).

While we observed site-variations in the factors associated with non-receipt of age-appropriate vaccination, child’s age and mother’s education were significantly associated with non-receipt of age-appropriate vaccination across six and three sites respectively (Table S1). Furthermore, we observed non-age appropriate vaccination to be less common in all study sites compared to Kenya but low mother’s education was positively associated with non-receipt of age-appropriate vaccinations in both infants and older children regardless of the site. Other factors observed in our study to be positively associated with non-receipt of age-appropriate vaccination included: low father’s education, dysentery and some dehydration among infants, and high heart rate and severe wasting among older children (≥ 12 months) (Table 4).

**Table 4:**
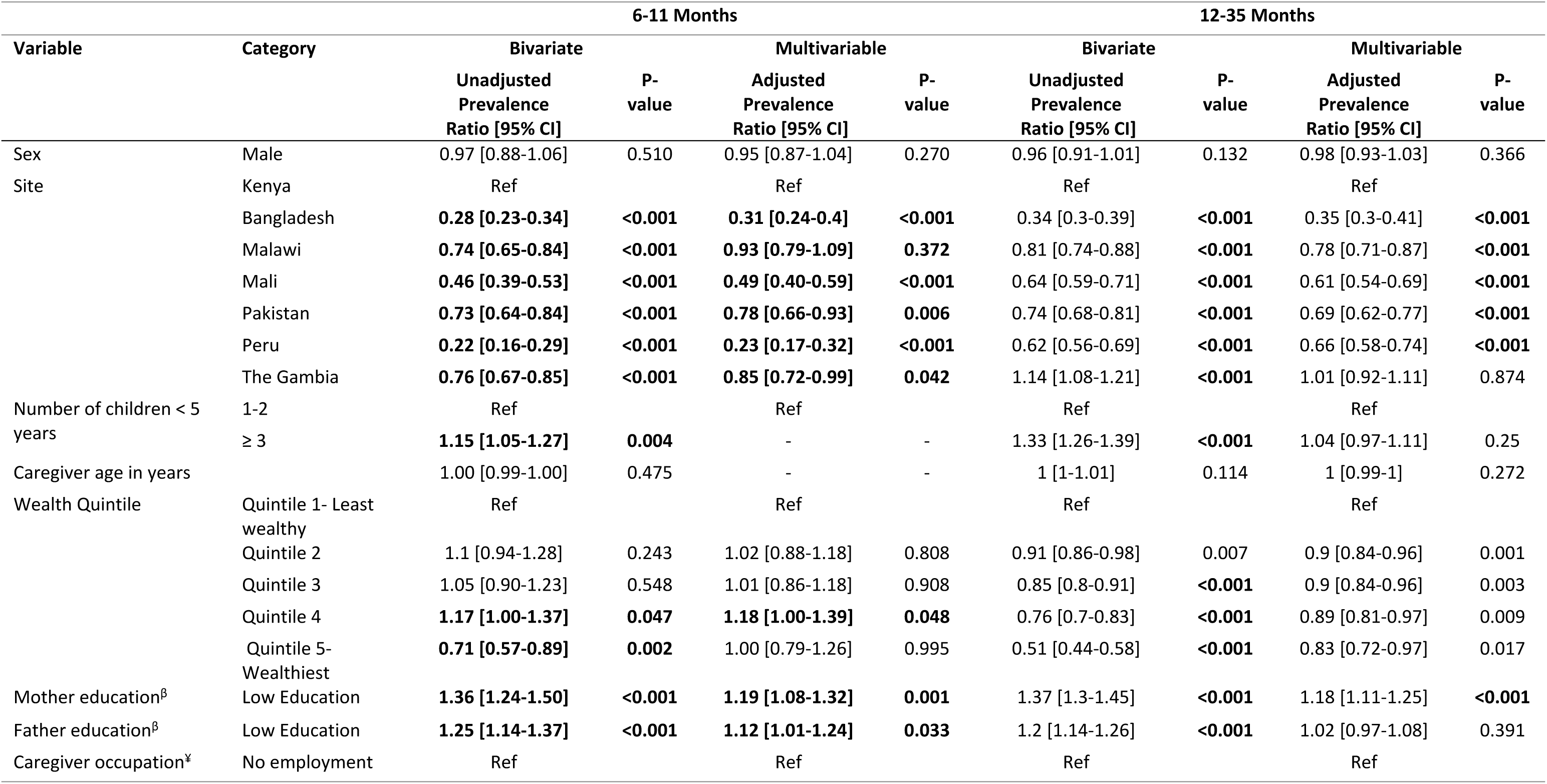
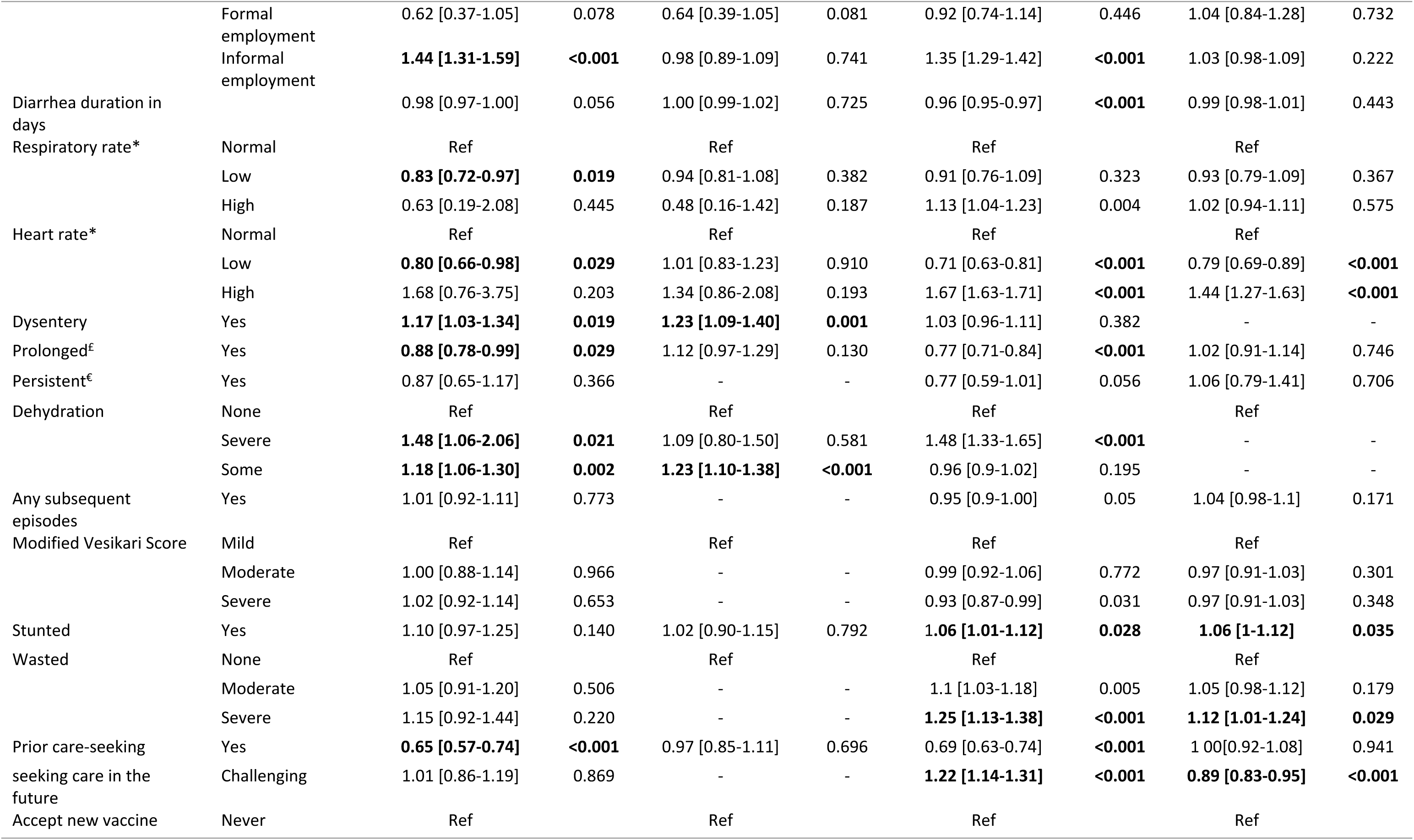
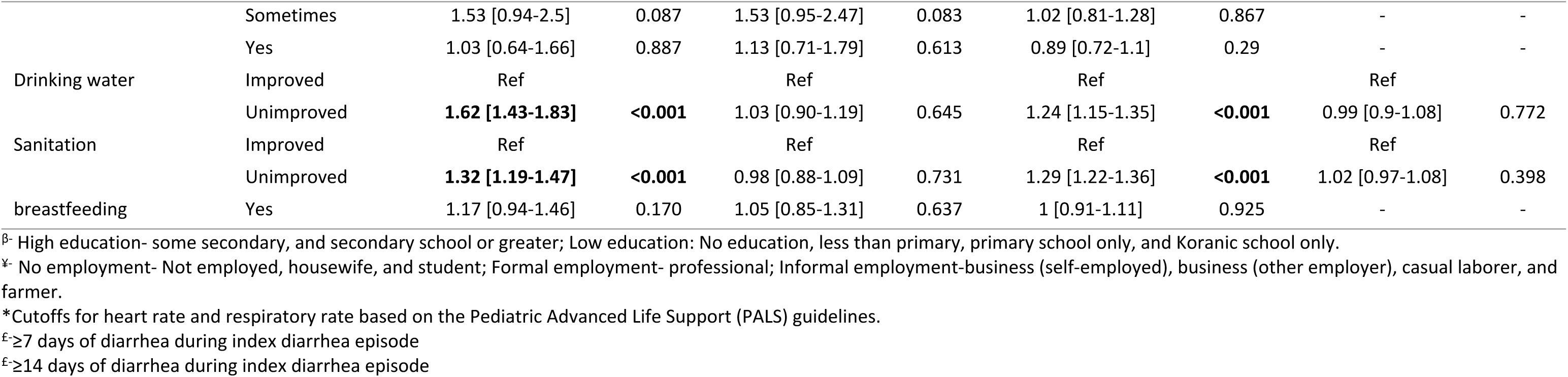
Age-stratified correlates of non-receipt of age-appropriate vaccination among children aged 6-35 months presenting with medically-attended diarrhea, EFGH *Shigella* surveillance study, June 2022 - August 2024.

## Discussion

More than half (52%) of the children enrolled in our study did not receive age-appropriate vaccination, with The Gambia reporting the highest prevalence (75.2%) and Bangladesh the lowest (22.3%). Vaccines with the highest prevalence of non-receipt of age-appropriate vaccination regardless of the site were: Polio (27%), MMR (19%) and PCV (14%) while BCG (3%) was the lowest, suggesting variations in missed opportunities for vaccination across the sites. Furthermore, predictors of non-receipt of age-appropriate vaccination included older age (24-35 months), severe wasting, caregivers taking care of three or more children <5 years in a household, low caretaker education and lower wealth index

Our findings suggest that more than half of the children enrolled in this study did not receive age-appropriate vaccination is important for several reasons. It suggests that a larger proportion of these children are inadequately protected against the intended vaccine preventable diseases, and that vaccine implementers in these settings should more than double their efforts in vaccinating children with Polio, MMR and PCV vaccines. This observation can possibly explain the persistently high infectious disease burden in these settings despite existing and routine vaccination programs. Our findings on low adherence for Polio, MMR and PCV are consistent with estimates from other recent studies showing that none of these countries may achieve the target for the third dose of Polio vaccine and measles at the regional level [18]. The reasons for delay in, or missing, the measles vaccine could include it being a single dose vaccine and that it targets children at a later age [19], consistent with observations from other resource poor settings [20]. The highest adherence to BCG vaccination is consistent with previous observations [18], and could be explained by an increasing trend in hospital births that usually coincides with the vaccine’s schedule [20–22].

Vaccination adherence declined with an increase in age, especially for vaccines scheduled later in childhood. This suggests caregivers’ domestic or socio-economic activities that may compete against taking their children for timely vaccination [23]. Worth noting from our study, is an observation that severe wasting, was associated with non-receipt of age appropriate vaccination, a finding that is consistent with literature from a systematic review by Favin et al [24].These findings may be explained in part by healthcare providers opting to defer vaccination in severely malnourished children due to safety concerns based on a perception that malnourished children may not tolerate vaccines well or may have reduced immune responses, leading to uncertainty about both safety and effectiveness as well as fear of adverse reactions [24,25]. Moreover, the association with wasting could also be explained by wasted children possibly coming from families who have less resources, less education, and overall tend to prioritize food and survival over health care attendance [26]. Additionally, three or more children under the same caregiver was also a driver of non-receipt of age-appropriate vaccination, with younger children being given priority over older children in such households, an observation which is consistent with some existing literature [5]. Our finding that children with MAD whose mothers reported lower education were more likely to have non-receipt of age-appropriate vaccination than their counterparts is in line with findings from other previous studies [27,28].

Children from relatively less wealthy or poor households were more likely to report non-receipt of age-appropriate vaccination, which is likely related to caregivers’ financial constrains which limit their free time due to competing economic engagements as a priority over taking their children for vaccination as observed in other previous studies [29,30]. Furthermore, caregivers with primary or less education were more likely to report a child with non-receipt of age-appropriate vaccination, possibly due to their lack of awareness of Expanded Programme on Immunization (EPI) schedules and associated benefits [31].

Our findings have several public health implications. First, maternal related factors are important drivers of age-appropriate vaccination in children from resource poor settings. Second, to optimize adherence to vaccine schedules, monitoring of context specific drivers of EPI schedules could improve vaccine uptake and enhance child survival strategies by targeting vulnerable children. Third, understanding factors associated with non-receipt of age-appropriate vaccination in LMICs could help inform resource allocation and strategies aimed at optimizing targeted individual vaccine uptake. Finally, should *Shigella* vaccine introduction target late infancy, then policy makers and vaccine implementers may need to consider innovative approaches to bolster its coverage given the observed EPI adherence challenges with vaccine administration in this age group [18].

Our study is subject to limitations. First, it was not powered to detect drivers of non-age-appropriate vaccination and vaccine promptness, hence our data needs to be interpreted with caution. Second, the impact of vaccine stock-outs, for instance stock-out of rotavirus vaccine in Kenya during the study period, was not evaluated in our study to help contextualize the results. Third, antenatal care data could complement and enhance understanding factors influencing receipt of age-appropriate vaccination in future studies. Finally, inconsistent adoption of vaccines into national EPI schedules across sites, such as Bangladesh not implementing rotavirus vaccination, may have limited our ability to comprehensively capture all relevant patterns of overall non-receipt vaccination assessment.

## Conclusion

Our study found that the prevalence of non-receipt of age-appropriate vaccination is significantly high across the sites. Given the variation, researchers need to carry out more studies to determine the potential causes of the variation by including additional explanatory variables, such as factors connected to health care services which can be valuable for policy makers in these settings.

From the policy perspective, our study highlights the critical role of engagement with health services and caregiver education in both improving the vaccination coverage as well as improving age appropriateness of the vaccinations.

## Data Availability

The EFGH statistical analysis plan (https://clinicaltrials.gov/study/NCT06047821) and study protocol (https://academic.oup.com/ofid/issue/11/Supplement_1) were made publicly available. The datasets were deidentified and anonymized and will be publicly available upon publication of the manuscript

https://academic.oup.com/ofid/issue/11/Supplement_1

## Acknowledgements

We are very grateful for the partners who made this work possible, including the ministries of health, health facilities and community partners of the various sites who worked tirelessly to support the implementation and data collection of the EFGH study. The EFGH study staff from each of the study sites worked together with absolute collaboration and enthusiasm to achieve the study objectives. We thank the EFGH Consortium and the Nyanja Health Institute who facilitated and contributed to the Manuscript Writing Cohort Program including Beth A. Tippett Barr and Erika Feutz for their support and guidance. Most importantly, we would like to thank the study participants and their families for their willingness to participate in this research study.

## Supplementary tables

**Table S1**: Site-specific correlates of non-receipt of age-appropriate vaccination among children aged 6-35 months presenting with medically-attended diarrhea, EFGH *Shigella* surveillance study, June 2022 - August 2024

